# Prevalence of plasmodium infection and associated risk factors among household members in Southern Ethiopia: Mult-site cross-sectional study

**DOI:** 10.1101/2023.11.22.23298901

**Authors:** Girma Yutura, Fekadu Massebo, Nigatu Eligo, Abena Kochora, Teklu Wegayehu

## Abstract

Despite continuous prevention and control strategies in place, malaria remains a major public health problem in sub-Saharan Africa including Ethiopia. This study is, therefore, aimed to determine the prevalence of plasmodium infection and associated risk factors in selected rural *kebeles* in southern Ethiopia. A community-based cross-sectional study was conducted between January and June 2019. Mult-stage sampling techniques were employed to select the study districts and *kebeles* from four zones. Blood sample were taken from 1674 participants by finger prick and thin and thick smears were examined by microscopy. Socio-demographic data as well as risk factors for malaria infection were collected using questionnaires. Bivariate and multivariate logistic regressions were used to analyze the data. The overall prevalence of malaria in the study localities was 4.5% (76/1674). The prevalence was varied among the study localities with high prevalence in Bashilo (14.6%; 33/226) followed by Mehal Korga (12.1%; 26/214). *Plasmodium falciparum* was the dominant parasite accounted for 65.8% (50/76), while *P. vivax* accounted 18.4% (14/76). Co-infection of *P. falciparum* and *P. vivax* was 15.8% (12/76). The prevalence of malaria was 7.8% (27/346) in age less than 5 years and 7.5% (40/531) in 5-14 years. The age groups >14years were less likely infected with plasmodium parasite (AOR=0.14, 95% CI 0.02-0.82). Asymptomatic individuals more likely had malaria infection (AOR = 28.4, 95% CI 011.4-70.6). Individuals living proximity to mosquito breeding sites have higher malaria infection (AOR = 6.17, 95% CI 2.66 - 14.3). Malaria remains a public health problem in the study localities with lower age group and asymptomatic individuals had higher plasmodium infection. Thus, malaria prevention and control strategies targeting children and asymptomatic cases are crucial to reduce malaria related morbidity and mortality.

## Introduction

Malaria is a life-threatening disease that causes morbidity and mortality worldwide. Globally, estimated 228 million cases 87 endemic countries reported in 2018 [1]. African regions particularly sub-Saharan Africa shares the highest burden of cases. The endeavor of malaria intervention strategies brought significant results in the past years. However, a total of 247 million cases reported in 2021 which is increased from 245 million in 2020 [2]. Between 2000 and 2019 the incidence of malaria cases decreased by 82 to 57 before increasing to 59 in 2020 [2]. Although malaria cases and deaths declined in last decades, there is slight increase in malaria cases and deaths and malaria remains a major public health problem putting approximately half world population at risk of infection [2, 3].

Five species of plasmodium namely *P. falciparum*, *P. vivax*, *P. ovale*, *P. malariae and P. knowlesi* cause disease in humans [4]. Of these, *P. falciparum* and *P. vivax* are distributed worldwide. *Plasmodium falciparum* infection dominates in Africa, while *P. vivax* is prevalent in Asia [5]. The two species are also the major causes of morbidity and mortality in Ethiopia. Despite the country’s long history of malaria control which dates back to the 1950s, the disease persistent to causes a significant threat [6].

Malaria transmission is often seasonal, variable, and bimodal in Ethiopia. It affects two-thirds of landmass, and more than 60 million people live in malaria-risk areas. *Plasmodium. falciparum* and *P. vivax* accounting to 60% and 40% of the disease in the country [7, 8]. *Plasmodium falciparum* is highly virulent species which causes severe malaria and death in the country [9, 10]. In Ethiopia there were 904,495 malaria cases and 213 deaths in 2019. *Plasmodium falciparum* caused most morbidity, with 724,996 (80%) followed by *P. vivax* 166,340 (18.4%) and 13,159 (1.5%), respectively [11].

The implementation of malaria preventive and control measures in Ethiopia, as in other areas of the world, has decreased malaria morbidity, death, and economic loss. In the fight against the disease, the distribution of long-lasting insecticide-treated bed nets (LLINs) and indoor residual spraying are critical. Additionally, increased healthcare utilization, early diagnosis, prompt treatment, prevention, rapid management of malaria epidemics, and disease surveillance were among the interventions used. Malaria prevention and control services have been provided free of charge in the country. Ethiopia is currently working on a malaria elimination program that aims to eradicate the disease by 2030 [12,13].

In the former South Nations Nationalities Peoples Regional State (SNNPRs) of Ethiopia, about 26% of the population is at risk and 65% of the area is malarious. The disease causes morbidity and mortality in the region [14]. The epidemiology of malaria is heavily influenced by altitude, season, climatic conditions, vector feeding behavior, socio-economic status, insecticide resistance status of vectors, antimalarial drug resistance status of plasmodium parasites, and the success of management methods. Hence, understanding the epidemiology of malaria at different geographical areas has public health implications. Therefore, the aim of this study was to assess the prevalence of malaria and the associated risk factors among communities in various geographical settings in selected sites of southern Ethiopia.

## Materials and methods

### Study areas

This study was conducted in four zones namely South Omo, Gamo, Wolaita, and Hadiya of the SNNPRs. The Zones were selected purposefully based on malaria report. Benatsemay, Kucha, Boreda, Humbo, and Misirak Badawacho districts were recruited from the selected zones. Of districts, eight *kebeles* (smallest administrative units in Ethiopia); namely: Enchete, Duma, Dana, Gocho Hambisa, Abaya Gurucho, Abaya Bilate, Mehal Korga, and Bashilo were selected (Fig 1). Average altitude of the study *kebeles* ranged from 553 m a. s. l at Duma to 1720m a. s. l. at Mehal Korga (Table 1). Malaria continues being a significant health problem in the region, but the transmission intensity varies across different local settings [15].

**Figure 1:**
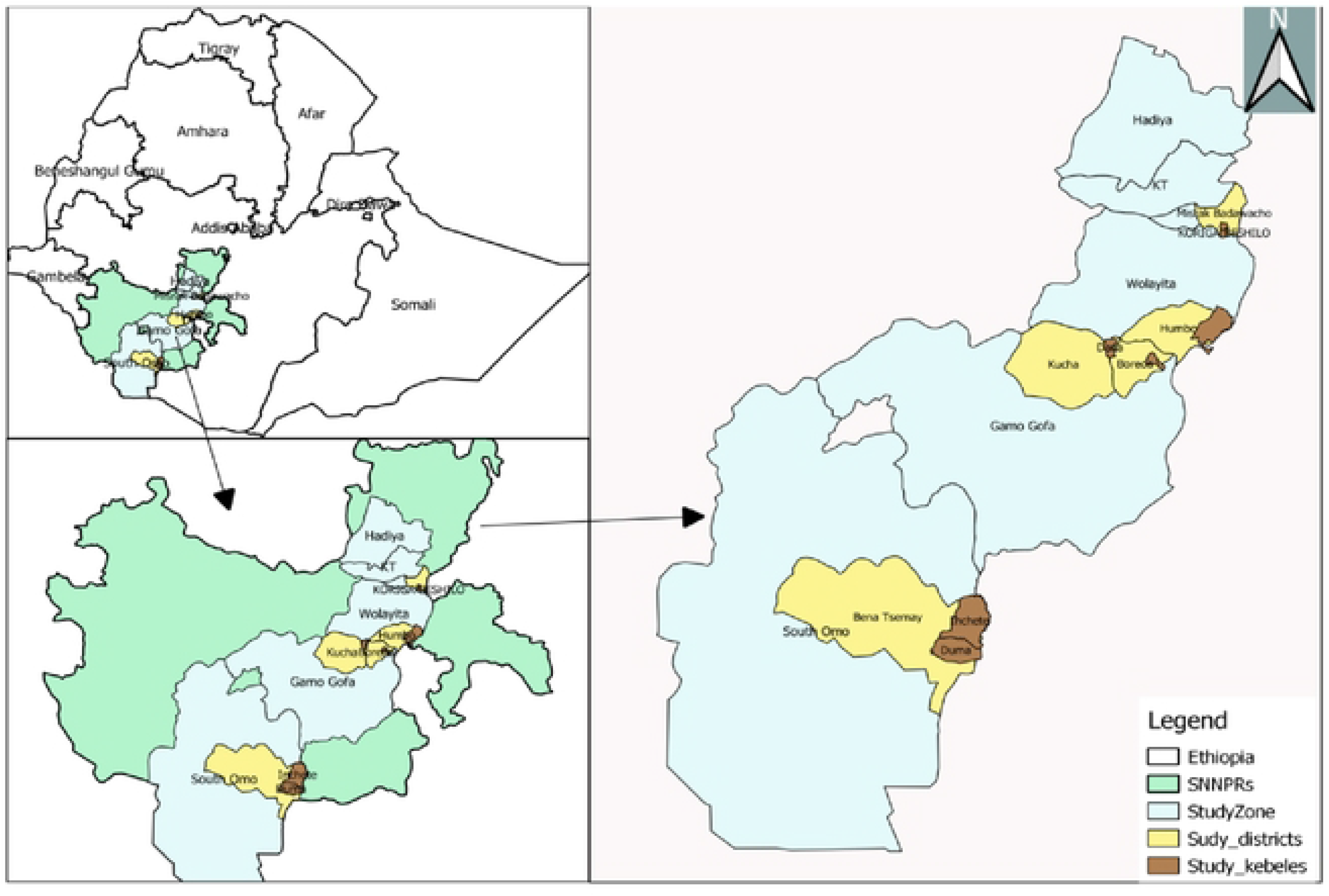
Map of Southern Nations Nationalities Regional State and study areas (made by Arc GIS version 10.1).

**Table 1:**
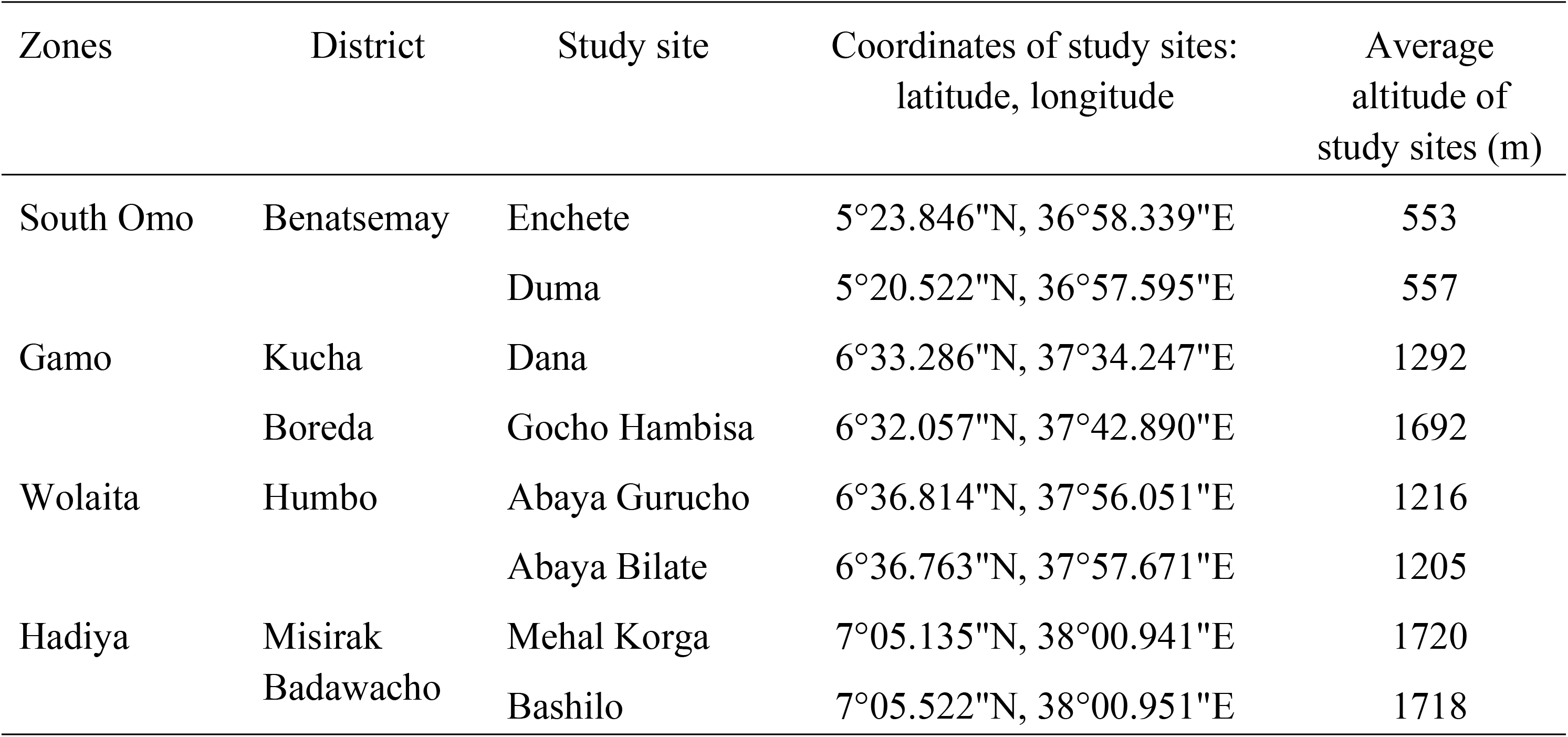
Geographical location of the study *kebeles* in four Zones in the southern Ethiopia, January-June, 2019.

### Study design

Community based cross-sectional study was conducted between January and June in 2019 to determine prevalence of malaria infection and associated risk factors among household members in randomly selected localities in southern Ethiopia.

### Study participants

People residing in all the study *kebeles* could be taken as source population and individuals in selected households were included as study participants based on the following inclusion and exclusion criteria.

### Inclusion and exclusion criteria

All household members regardless of the age groups and sex who lived in the *Kebele* for at least 6 months were included in the study. Individuals, who receiving malaria treatment during survey as well as non-consenting respondents were excluded.

### Sample size determination and sampling techniques

The sample size was determined using single population proportion formula of Fink and Kosecoff [16] assuming, 16% expected prevalence [17], 2.5% margin error, design effect 2, α=5% (95% confidence level), and 15% non-response rate. Accordingly, the calculated sample size was 1,674 individuals. From the total sample size, the study participants were allocated proportionally based on entire population for each study site.

Multistage sampling technique was employed to sample study participants in the districts. According to Ethiopian population and housing census of 2007, average family size for the region ^was 4.9^ [18]. Hence, the 1,674 individuals were calculated based on their population size and 342 _(1674/4.9)_ households were in 8 rural *kebeles* (Table 2).

**Table 2:** Sample size allocation to malaria prevalence study *kebeles* in four Zones in the southern Ethiopia, January-June, 2019.

| **Zones** | ***Kebeles*** | **Total population** | **HH (Pop/ 4.9** | **Sample size allocation** | **No. of sampled HHs** |
| --- | --- | --- | --- | --- | --- |
| South Omo | Enchete | 1934 | 395 | 140 | 29 |
|  | Duma | 1607 | 328 | 116 | 24 |
| Gamo | Dana | 2980 | 608 | 216 | 44 |
|  | Gocho Hambisa | 3232 | 660 | 234 | 49 |
| Wolaita | Abaya Gurucho | 3357 | 685 | 243 | 50 |
|  | Abaya Bilate | 3351 | 684 | 243 | 50 |
| Hadiya | Mehal Korga | 3136 | 640 | 224 | 46 |
|  | Bashilo | 3575 | 730 | 259 | 53 |
| **Total** |  | **23172** | **4729** | **1674** | **344** |

Key: HH =household

These households were included by employing a systematic random sampling technique, during sample collection, to obtain individual participants. The lists of households from health posts were used as sampling frames with an assumption of similar exposure to vector control interventions. The first household was selected randomly by lottery method and every k^th^ household was included in the study. Where K is calculated by the formula of 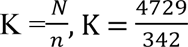, Where, K= the gap between every household, N=total number of households in the study *kebeles* and n=sample size of households was calculated from individual sample size (Table 2). Therefore, K = 13, thus, every 13^th^house hold was used to obtain individuals participants in the study *kebeles*. Each household was coordinated using geographical position system for geographical location.

### Sample collection and processing

#### Blood sample collection and processing

Capillary blood sample was collected using sterile blood lancets from participants after obtaining consent during house-to-house visits. Blood sample collection was done by senior medical laboratory technicians, following standard guidelines [19]. Air dried blood smears were transported to nearby health centers’ laboratories using slide boxes. The smears were fixed using 99.8% methanol, dried, and stained with a 10% Giemsa solution for 15 minutes. Then, microscopy was employed by experienced laboratory technicians to detect and identify plasmodium parasite species according to laboratory guidelines.

#### Socio-demographic data collection

Socio-demographic data were collected from 342 households based on structured questioner. The questioner prepared in local language was sought information on socio-demographic characteristics, and malaria prevention and control practices. After having the verbal consents, both individual and household-level factors associated with malaria transmission was obtained from the participant. During the time of sample collection, fever of study participants was checked and signs and symptoms of malaria such as headache, chills, sweating were asked. Fever of individuals was measured using thermometers.

### Data quality assurance

Data quality was maintained using various approaches. First, training was given for field assistants (data collectors) to have a common understanding to collect the appropriate demographic information. Second, blood sample collection and microscopy were done by senior laboratory technologists and discussion was held to apply standard operational diagnostic procedures during laboratory work. Each questioner and the collected sample was cross-checked for completeness, accuracy, and consistency by the group members and corrective measures taken. Moreover, all houses were coordinated using geographical position system and study individuals were coded during blood sample collection. All positive slides and 10% of negative slides were re-examined by another senior laboratory technologist blinded to previous slide results. Slides were declared negative for plasmodium parasites after thorough examination of 100 fields and no plasmodium parasite is detected by microscopy.

### Study variables

The outcome variable for examination of blood films was binary variables and indicated as positive or negative for plasmodium parasites. Independent variables included house structure (the roof material, floor material, presence of visible holes on wall), IRS spraying in the last 12 months, LLINs ownership (presence of bed nets, total number of nets, access to LLINs and use of mosquito nets), presence of mosquito breeding site. The variables like sex, age, educational status and fever (auxiliary temperature) were considered as individual level for analysis of data.

### Data analysis

Data was entered into Microsoft Excel spreadsheets and analyzed using SPSS version 20.0. Descriptive statistics were used to determine the frequencies of variables. Bivariate logistic regression analysis was conducted to examine the association between plasmodium infections with associated risk factors. Multivariate logistic regression analysis was conducted to test potential predicators’ variable that was the main risk factor for malaria infection. The goodness of model fit was checked by Hosmer-Leme show-test and the logistic regression was fit for the test. Data normality was checked by non-parametric test of one-sample Kolmogorove-Smirnov test (1-sample K-S). During binary logistic regression if the *p* ≤ 0.025 was considered as a candidate for multivariate logistic regression. Thus, logistic regression statistical method of multivariate logistic regression was used with a 95% confidence interval and odds ratio was used to control confounders with the level of statistical significance was taken as *P*-value <0.05 for analysis of independent and outcome variables.

### Ethics approval

The study was reviewed and approved by the Ethical Review Committee of Arba Minch University (Ref.No.CMHS/12033592/111). Prior to the study, permission letter was obtained from selected Zonal Health Departments. Consent was obtained and official letter was taken from the respective district’s health office of the study *kebeles* administrators. The purpose of the study and procedure of blood sample collection were explained to the participants. The participants’ written and verbal consents were obtained from their parents/guardians of children and younger. The Study participants those positive for *P. falciparum* and *P. vivax* was treated free of charge at nearby health facilities.

## Results

### Socio-demographic characteristics of the participants

The socio-demographic characteristic of the study participants was summarized in Table 3. From the total of 1674 participants, 748 (44.1%) were males and 926 (54.5%) were female. With regard to the age, 346 (20.4%), 531 (31.7%) and 797 (44.5%) were in the age groups < 5, 5-14 and >14 age groups, respectively. Of the total, 1638 (97.8%) were asymptomatic and the rest 13 (2.2%) were symptomatic cases.

**Table 3:**
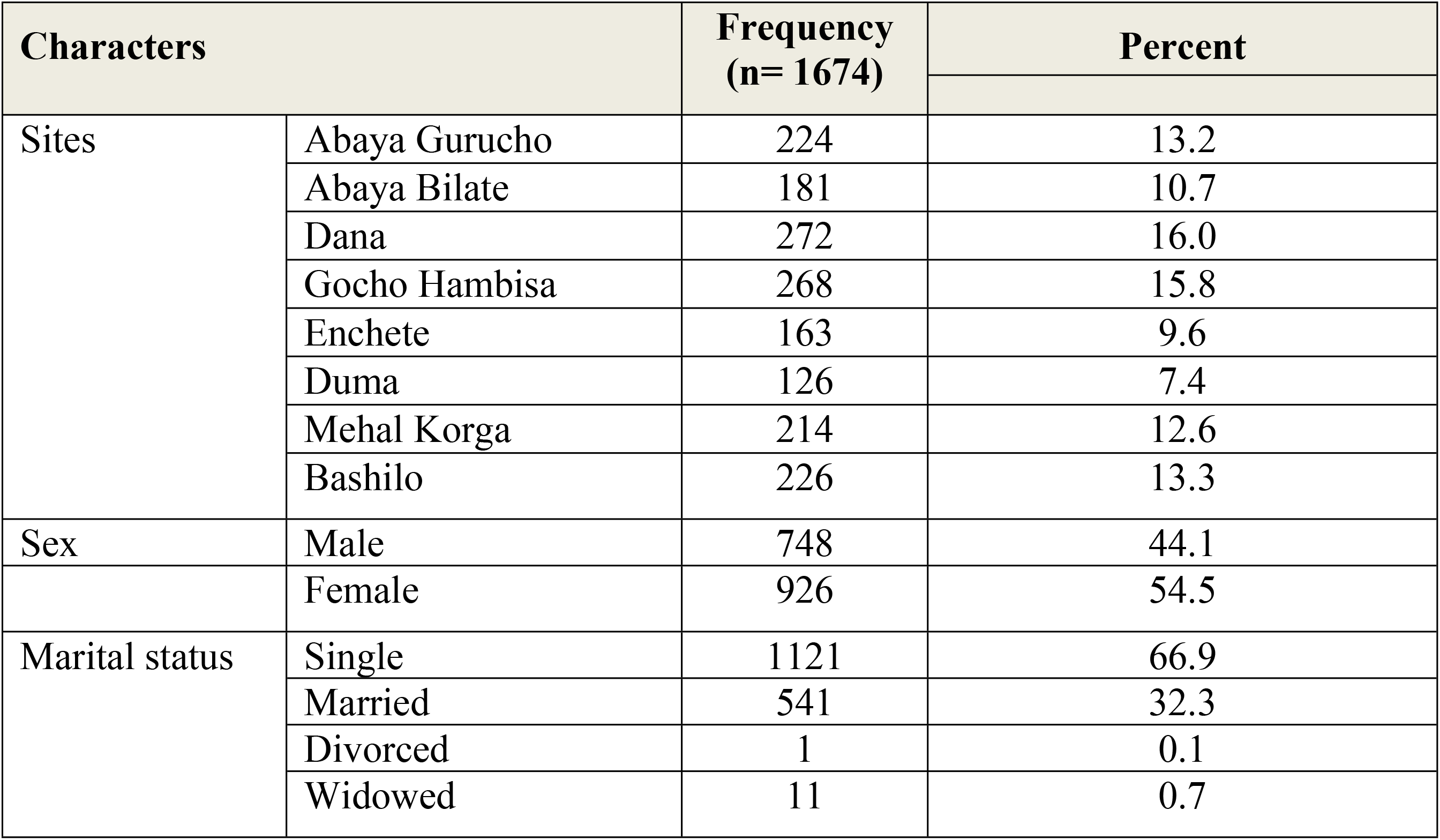
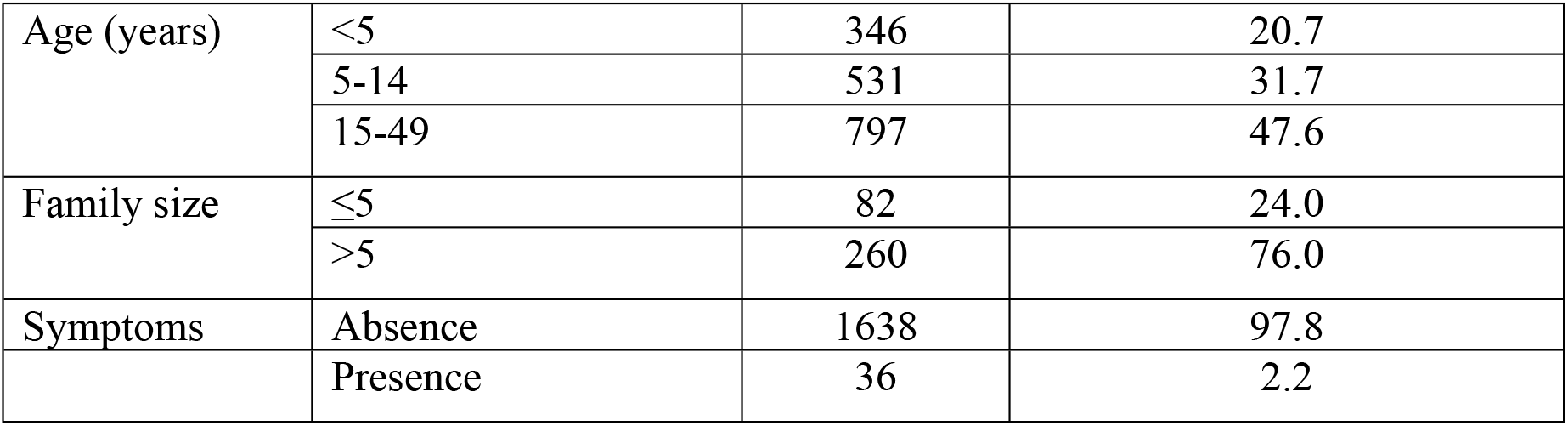
Socio-demographic characteristic of study participants by study *kebeles* in the southern Ethiopia, January-June 2019.

### Prevalence of malaria

The overall prevalence of malaria was 4.5% (76/1674) confirmed by microscopy (Table 4). The plasmodium infection was more prevalent in Bashilo *kebele* 14.6% (33/226) followed by Mehal Korga 12.1% (26/214). Plasmodium infection was detected in seven study *kebeles* and no malaria cases were detected in Gocho Hambisa *kebele*.

**Table 4:**
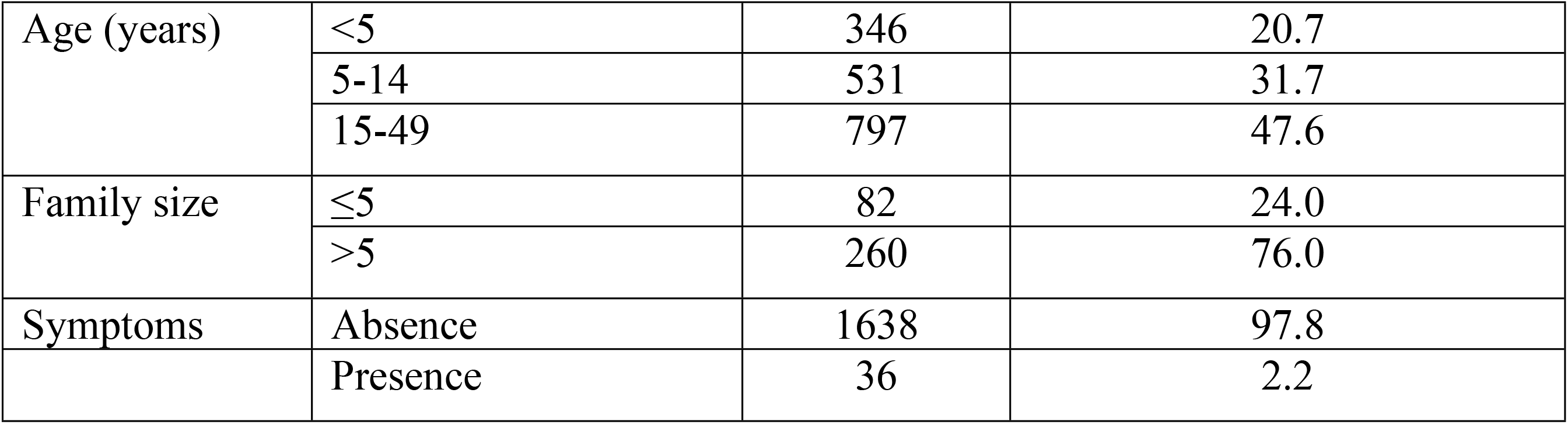
Prevalence of plasmodium species in eight *kebeles* of the southern Ethiopia, January-June 2019.

### Identification of plasmodium species

Among the confirmed malaria cases, *P. falciparum* was dominant species accounting 65.8% (50/76), while *P. vivax* was 18.4% (14/76). Mixed infections with *P. falciparum* and *P. vivax* were accounted 15.8% (12/76). Higher prevalence of *P. falciparum* 10.18% (23/226) was observed in Bashilo *kebele*. Among study *kebeles*, Mehal Korga had the high prevalence of *P. vivax* 3.74% (8/214) (Table 4).

### Sex and age-related prevalence of malaria

Of the study participants, 5.2% (39/748) males and 4% (37/926) females were found positive for plasmodium parasite (Table 5). The prevalence of plasmodium parasites among age groups were 7.8% (27/346) in under five children, 7.5% (40/531) in 5-14 years and 1.1 (9/797) in >14 years. The greatest malaria prevalence was observed among under five children followed by school age groups.

**Table 5:** Sex and age-related prevalence of malaria in study setting of the southern Ethiopia, January-June 2019.

| Variables |  | Study Participants | Malaria positive cases | Percentage (%) |
| --- | --- | --- | --- | --- |
| Sex | Male | 748 | 39 | 5.2 |
|  | Female | 926 | 37 | 4.0 |
| Age | <5 | 346 | 27 | 7.8 |
|  | 5-14 | 531 | 40 | 7.5 |
|  | >14 | 797 | 9 | 1.1 |

### Associated risk factors for malaria

A total of nine independent variables were considered for bivariate logistic regression analysis of individuals and household associated risk factors for plasmodium parasite infections (Table 6). The variables associated with individual and household-level risk factors of plasmodium parasite infection were age, fever during survey time, marital status, LLINs utilization, IRS spray status, house structure (main roof material), main wall material, presence of visible hole on the wall, and living proximity to breeding sites. Among those variables, the age of individuals, marital status, fever, LLINs utilization and living proximity to the breeding site were a candidate for multivariate analysis.

**Table 6:** Risk factors of plasmodium infection in the study localities in southern Ethiopia, January and June, 2019.

| Variables |  | Category |  | Binary logistic |  |  | Multivariate logistic |  |  |
| --- | --- | --- | --- | --- | --- | --- | --- | --- | --- |
|  |  | +ve(%) | -ve (%) | P-value | COR | CI | P-value | AOR | CI |
| Age | <5 | 27(7.8) | 319(92.2) |  |  |  |  |  |  |
|  | 5-14 | 40(7.0) | 532(92.5) | 0.89 | 0.97 | 0.58-1.60 | 0.83 | 1.08 | 0.54-2.14 |
|  | >14 | 9(1.2) | 747(98.9) | 0.001* | 0.14 | 0.07-0.31 | 0.029* | 0.14 | 0.02-0.82 |
| Marital Status | Single | 68(89.5) | 1065(66.6) | 0.001* | 0.24 | 0.11-0.49 | 0.51 | 1.83 | 0.3-11.9 |
|  | Married | 8(10.5) | 533(33.4) |  |  |  |  |  |  |
| Fever | No | 53(3.2) | 1585(96.8) |  |  |  |  |  |  |
|  | Yes | 23(63.9) | 13(36.1) | 0.001* | 53.93 | 25.9-112.4 | 0.001* | 28.4 | 11.4-70.6 |
| LLINs Utilization | No | 39(7.4) | 487(92.8) |  |  |  |  |  |  |
|  | Yes | 37(3.2) | 1111(96.8) | 0.001* | 0.43 | 0.27-0.68 | 0.003* | 0.37 | 0.19-0.72 |
| IRS Spray | No | 46(5.2) | 837(94.8) | 0.166 | 0.72 | 0.45-1.15 | 0.06 | 0.52 | 0.26-1.03 |
|  | Yes | 30 (3.8) | 761(96.2) |  |  |  |  |  |  |
| Roofing | Corrugated | 28 (3.8) | 710(96.2) | 0.195 | 1.37 | 0.85-2.21 | 0.23 | 0.66 | 0.33-1.31 |
|  | Thatched | 48 (5.1) | 888(94.9) |  |  |  |  |  |  |
| Wall | Wood | 5 (1.4) | 290(98.6) | 0.89 | 0.86 | 0.09-7.78 | 0.47 | 0.40 | 0.03-4.78 |
|  | Wood and mud | 70 (5.4) | 1246(94.6) | 0.21 | 3.53 | 0.48-25.8 | 0.39 | 2.46 | 0.31-19.3 |
|  | Wood cement | 1(1.6) | 62(8.4) | 1 |  |  |  |  |  |
| Visible wall hole | No | 42(5.4) | 733(94.6) |  |  |  |  |  |  |
|  | Yes | 34(3.8) | 861(96.2) | 0.12 | 0.69 | 0.43-1.10 | 0.63 | 0.85 | 0.44-1.64 |
| Breeding site | No | 12(2.0) | 578(98.0) |  |  |  |  |  |  |
|  | Yes | 64(5.9) | 1020(94.1) | 0.005* | 2.6 | 1.34-5.05 | 0.001* | 6.17 | 2.66-14.3 |

In the multivariate logistic regression analysis, the predictors of plasmodium infections after controlling confounders of the variables were the age of individuals (AOR=0.14, 95% CI 0.02-0.82) and fever during survey time (AOR= 0.37, 95% CI0.19-0.72). Household-level predictor variables of plasmodium infections were LLINs utilization (AOR=0.37, 95% CI 0.19-0.72) and proximity mosquito breeding sites (AOR=6.17, 95% CI 2.66-14.3) were a significant association with plasmodium infection.

The individuals who’s aged <5 was 86% more likely to have a malaria as compared with individuals whose age >14 with the p-value =0.029 (IC=0.02-.82). Individuals who do not have a fever during study time were 28.4 times more likely have plasmodium parasite as compared to individuals with fever with the p-value=0.001 (CI=11.4-70.06).

LLINs utilization was significantly associated with plasmodium species. The individuals that have not to use LLINs during a sleeping time were 63% more likely have a chance to plasmodium parasite infection as compared with their counterparts with the p-value=0.003 (CI=0.19-0.72) (Table 6). Those individuals who live proximity to the breeding site were 6.17 times more likely have a chance to develop malaria as compared to individuals do not live around breeding site with the p-value=0.001 (CI=2.66-14.3).

## Discussion

Malaria affects the lives of almost all people living in sub-Saharan African countries. In Ethiopia, malaria remains a major public health problem despite continuous control and preventive strategies in place. The overall prevalence of malaria infection in this study was 4.5% with varying prevalence in different study sites in southern Ethiopia. Both *P. falciparum* and *P. vivax* has been identified with *P. falciparum* dominant species accounted for 65.8% (50/76). It was also observed that lower age group, asymptomatic case, individuals who live proximity to mosquito breeding site had higher plasmodium infection.

The overall prevalence of plasmodium infection in this study (4.5%) was in line with reports from various parts of Ethiopia including 4.4% in Butajira, 6.1% in Benatsemay district (South Omo), 6.7% in Dembia districts, 6.8% in Sanja town, and 4% in Jimma zone [20–23]. This finding is higher than the prevalence reported in another study in Butajira and national malaria indicator survey 2015 result, with prevalence of 0.9% and 0.5%, respectively [24, 25]. On the other hand, the present finding is much lower than the prevalence reported in Kisumu country in the Kenya with 28% [26], Armachiho districts, North West Ethiopia with 18.4% [27], and Dilla town and surrounding areas with 16.0% [17]. The difference in findings might be associated with sociodemographic, socioeconomic and environmental factors that could affect the epidemiology of malaria such as altitude, proximity to mosquito breeding sites, housing condition, ownership and utilization of LLINs among others.

Prevalence of malaria infection was relatively high in Bashilo (14.6%) and Mehal Korga *kebeles* (12.6%) were observed compared to other study areas. The study conducted different parts in Ethiopia showed that the prevalence of plasmodium infection was different among study participants [20, 28, 29]. In the study areas, heterogeneity of plasmodium infection in the study settings in the local community was existed. The reason is why because of ecologic and environmental factors, host and vector characteristics, social, biological and socio-demographic factors in the study areas.

*Plasmodium falciparum* and *P. vivax* were identified as co-endemic species in study areas while *P. falciparum* was dominant species of parasite. The dominance of *P. falciparum* was consistent with the study conducted in Benatsemay districts in south Omo, Ethiopia [23]. In addition, the national community-based malaria indicator surveys conducted during peak malaria transmission season in the 2007 and 2011 reported the dominance of *P. falciparum* as 83% and 77%, respectively [30,31]. The dominance of *P. falciparum* species might be more widely distributed in many parts of Ethiopia. This might be associated to the capacity of *P. falciparum* parasite to develop resistance against anti-malarial drugs represents a central challenge in the global control and elimination of malaria [32]

In contrast to this finding, other studies conducted in different geographical settings in Ethiopia [28,29] monitoring changing of the epidemiology of malaria beyond Gark projects [33] and the facility-based cross-sectional study in Hadiya Zone [34] the *P. vivax* dominates over *P. falciparum*. One possible reason for predominance of *P. vivax* might be improper management of primaquine that lead to the relapse of hyponozoites.

Regarding the age groups, the likelihood of having higher malaria cases was found among under five children and school age children than other age groups. The finding of this line with malaria prevalence in Ethiopian malaria indicator survey [25], the study conducted in Arba Minch Zuria district [35] children this age groups are more vulnerable and had have plasmodium parasite infections. The reason why high malaria cases in the age groups, the children of these age groups are somewhat more risk groups due to immunity status, exposed to mosquito bites, and less attention of care for utilization of malaria preventive measures.

Asymptomatic malaria infection was common in endemic areas. In malaria-endemic areas, people may develop partial immunity, allowing the asymptomatic infection to occur. The odds of plasmodium infection were higher on individuals those do not show symptoms (fevers) than the symptomatic cases in the study site. The result consistent with the study conducted in Senegal that indicated *P. falciparum* was dominant species in asymptomatic cases [36]. In other way, in low transmission settings, asymptomatic cases are common and most of the asymptomatic infections are sub-microscopic [28,37]. Study showed that asymptomatic cases could serve as reservoirs of infections to the mosquito vectors [38], Thus, they could serve as a major source of gametocytes and contributed to residual transmissions of malaria as asymptomatic carriers do not visit health facility for treatment. In many countries *P. falciparum* is asymptomatic or sub-clinical. In very low transmission settings, sub-microscopic carriers may contribute up to 50 % of humans to mosquito transmission [39].

Appropriate use of the utilization of LLINs is one of the key interventions for the prevention of malaria [40]. In this study, ownership of LLINs was 76.9%. This finding was higher than the previous findings in Hadiya zones with LLINs ownership of 41.6% [34]. On the other hand, national malaria indicator survey conducted in 2011 and 2015 showed 55 % and 64% of households have at least one LLINs of any type [25,30] and a community-based cohort study in South Central Ethiopia [41]. However, the accesses to LLINs were not significantly associated with plasmodium infection in study sites.

The utilizations of LLINs have an association with malaria cases among study participants. The current study showed that participants who use LLINs had lower malaria cases than those do not use. This findings in line with the study conducted Dilla and surroundings areas, Dembia districts, and Hadiya zones where participants do not use bed nets were 0.2, 0.2 and 4.6 times more likely developed plasmodium parasite infections, respectively [17,22,34]. The finding speculates the proper usage of LLINs protects from malaria through protecting mosquito bites depending on biting activity. It is noticeable that the proper utilization of LLINs will prevent mosquito that in turn prevent plasmodium parasite infection. These findings might the implication of possession and efficacy of LLINs utilization in the community and less attention to frequent utilization in different local settings.

Another important factor that determines the odds of plasmodium infection is living proximity to the breeding site. In this study, a participant who live proximity to mosquito breeding sites was at high risk of plasmodium infections. The study participants those lives proximity to the stagnant water of mosquito the breeding sites 6.17 times more likely have a chance to develop plasmodium infection as compared to individuals do not live around the breeding site. This finding in agreement with the study conducted in Dilla and surrounding areas and Dembia districts [17,22] by increasing the probability of having plasmodium infection. This is because proximity mosquito breeding sites give more chances to exposure mosquito bites in the community. It is fact that environmental management is important measures to prevent mosquito breeding sites to reduce malaria transmission.

Our study has some limitations. One of the limitations of this study is the laboratory diagnosis which is limited to microscopy only, a low sensitive tool. We were not able to determine the temporal relationship of disease as the study design is cross-sectional. The community-based nature of the study can be viewed as one of the strengths of this study as it enables us to screen the asymptomatic cases who could serve as potential reservoir of malaria parasite. High response rate of study participants can also be viewed as another strength of this study.

## Conclusions

Malaria is still important public health problems, although the prevalence of disease was varying in the study sites. Lower age children were at higher risk of plasmodium infection and more asymptomatic cases. Thus, malaria prevention and control strategies addressing lower age children and asymptomatic cases should be in place to reduce malaria associated morbidity and mortality in the study localities.

## Data Availability

All relevant data are within the manuscript and its Supporting Information files.

## Competing interest

no interest

## Author’s contributions

Conceptualization the gap: Girma Yutura, Fekadu Massebo, Nigatu Eligo, Teklu Wegayehu

Data accusation and management: GirmaYutura, Nigatu Eligo

Data analysis and write up: Girma Yutura, Fekadu Massebo, Abena Kochora, Teklu Wegayehu

Review and editing: Girma Yutura, Fekadu Massebo, Teklu Wegayehu

## Acknowledgements

We would like to thank all the health centers and laboratory technicians of study sites for their cooperation during sample collection and processing. We are also grateful to South Omo, Gamo, Wolaita and Hadiya zonal and districts health departments and *Kebele* administrators for their technical support. Study participants are acknowledged for taking parts in the study and we would like to thank Arba Minch University for its financial and technical support.

## Notes

### Competing Interest Statement

The authors have declared no competing interest.

### Funding Statement

Yes

### Author Declarations

The study was reviewed and approved by the Ethical Review Committee of Arba Minch University (Ref.No.CMHS/12033592/111).

